# Digital Health Technologies for Metabolic Disorders in Older Adults: A Scoping Review Protocol

**DOI:** 10.1101/2024.02.26.24303372

**Authors:** Panitda Huynh, Elgar Fleisch, Michael Brändle, Tobias Kowatsch, Mia Jovanova

## Abstract

**Introduction:** Metabolic disorders (type 2 diabetes, insulin resistance, hyperglycemia, obesity, hyperlipidemia, hypertension, nonalcoholic fatty liver disease, and metabolic syndrome) are leading causes of mortality and disability worldwide and disproportionately affect older adults relative to those younger. Digital health technologies (DHTs), such as patient monitoring, digital diagnostics, and digital therapeutics, emerge as promising tools for navigating health in day-to-day life. However, their role in targeting metabolic disorders, particularly among a key demographic of older adults, is not yet fully understood. Thus, this work aims to scope the use of DHTs in managing metabolic health disorders among older adults.

**Methods and Analysis:** We will conduct a scoping review following the recommended framework by Arksey and O’Malley (1). Our search will focus on three primary concepts: metabolic disorders, DHTs, and older adults. We plan to search five online databases—Cochrane, Embase, PubMed, Scopus, and Web of Science—to identify original research articles published between January 2014 and January 2024. Two reviewers will independently screen articles for inclusion based on predetermined criteria, and a separate reviewer will resolve conflicts. Data will be extracted using a standardized form, and the findings will be synthesized and reported qualitatively and quantitatively.

**Ethics and dissemination:** No ethics approval is required for this protocol and scoping review, as data will be used only from published studies with appropriate ethics approval. Results will be disseminated in a peer-reviewed publication.

This protocol has been preregistered on OSF Repository at: https://osf.io/9s8fm.

**Strengths and limitations of this study:**

- To our knowledge, this scoping review is the first to scope the landscape of DHTs for targeting metabolic disorders among older adults.
- We apply the DHT Ecosystem Categorization for a more standardized overview of the peer-reviewed empirical literature across multiple databases and follow rigorous scoping review reporting guidelines.
- Consistent with the nature of scoping reviews, our study does not include an assessment of the quality of the included studies, only involves studies in English, and does not include non-peer reviewed industry reports, which may also provide relevant information.
- Since our pre-existing definition of metabolic disorders focuses on various pre-defined major conditions, there exists a possibility that we may not comprehensively capture all possible instances of metabolic disorders among older adults.

## 1 BACKGROUND

Non-communicable diseases (NCDs) are primary drivers of morbidity and mortality globally, with estimates by the World Health Organization (WHO) to surpass 15 million premature deaths attributed to NCDs per year (2). Over 50% of cases of NCDs is attributed to a growing prevalence of metabolic disorders; namely type 2 diabetes mellitus (T2DM), hypertension (HTN), hyperlipidaemia (HLD), obesity and more recently, non-alcoholic fatty liver disease (NAFLD) (2,3). Metabolic disorders are more prevalent among older adults versus younger populations, in part due to age-related physiological changes in metabolism and hormones, cumulative effects of lifestyle factors (e.g., nutrition, sleep, and physical activity), and susceptibility to related comorbidities (e.g. cardiovascular diseases, diabetes) (4–6). Metabolic disorders are often defined as conditions that affect the body’s conversion of food into energy and the ability to eliminate waste (7). Key factors contributing to the risk of metabolic disorders are the intake of an unhealthy diet, a sedentary lifestyle, and insufficient exercise, among others (7).

Nonpharmaceutical intervention efforts that target lifestyle factors are a promising approach to navigating metabolic disorders (7), and increasingly involve digital health technologies (DHTs)– which are defined by the Food and Drug Administration (FDA) as “computing platforms, connectivity, software, and sensors used for health care and related uses (8,9). Notably, DHTs may help older adults and caregivers manage aspects of metabolic health in day-to-day life (vs. in the clinic) and overcome barriers of more traditional health assessments (10), e.g., by reducing costs associated with in-person visits for patients and reducing burden for healthcare professionals, enhanced experience for patients, as well as improved mental health for healthcare professionals (11). However, DHTs are rapidly evolving and are often studied under inconsistent terminologies (10,12,13). This heterogeneity complicates efforts to synthesize evidence on their prevalence and potential impact in metabolic and related health contexts (13,14).

In response, the international organization overviewing DHT implementation in healthcare, the Digital Therapeutics (DTx) Alliance, has called for systematic overviews of DHTs and a shared language to facilitate standardization (8,9). To this end, DTx Alliance has formally categorized DHTs according to their intended purpose in five categories (see Table 1): (i) promoting health & wellness, (ii) patient monitoring, (iii) care support, (iv) digital diagnostics, and (v) digital therapeutics (15). These classification guidelines aim to help map a wide range of existing products by their intended use and offer a more consistent and actionable language for different stakeholders, including researchers, medical professionals, patients, and policymakers (8).

**Table 1:** Digital Health Technology Ecosystem Categorization.

| **DHT Category** | **Health & Wellness** | **Patient Monitoring** | **Care Support** | **Digital Diagnostics** | **Digital Therapeutics** |
| --- | --- | --- | --- | --- | --- |
| Definition | Disease-agnostic digital health solutions that primarily capture and store general health data and promote healthy living | Digital solutions intended to monitor specific health data, which may be interpreted by physician for clinical management | Digital solutions intended to help patients better manage their care of a specific disease or medical condition | Validated digital tools and software that deliver a diagnosis or prognosis of a specific disease or medical condition | Health software intended to treat or alleviate a specific disease or medical condition by generating and delivering a medical intervention |
| Claims | No claims to treat, improve, or diagnose a | May make non-clinical claims to assess patient | May make non-clinical claims to improving | Make a clinical claim to diagnose or assess a | Make a clinical claim to treat or alleviate a spe- |
|  | medical condition | data | health adjacent measures (e.g., adherence) | specific disease or medical condition | cific disease or medical condition |

Note: The Digital Health Technology Ecosystem Categorization is adapted from DTx Alliance and provides a uniform overview of different DHT categories by their intended use. (8)

In line with recent calls for systematic synthesis of DHTs in healthcare (13,16–18), several scoping reviews have recently overviewed the role of digital health applications in navigating NCDs (14,19–21), health promotion (22), and their associated barriers and facilitators (19–21) among older adults. In parallel, a separate line of work has begun to review the scope of interventions in promoting behaviors related to metabolic health, such as physical activity and nutrition among older adults (5,23). However, this work has not yet considered specific the role of DHTs in the context of metabolic health. Bridging these disparate lines of work, we aim to scope DHTs in metabolic disorders among older adults, encompassing a twofold objective. First, following guidelines from the DTx Alliance, we overview the prevalence of DHTs, and different DHT sub-categories, in navigating metabolic disorders in older adults. Second, we seek to identify how are DHTs used to address metabolic disorders among older adults and by whom. To our knowledge, this review will be the first to provide an overview of the DHT landscape in metabolic disorders among older adults.

## 2 METHODS AND ANALYSIS

For this review, we will adopt the framework outlined by Arksey and O’Malley (1), which involves a five-stage process: (i) formulating the research question, (ii) locating relevant studies, (iii) choosing studies that meet our criteria, (iv) extracting and organizing data, and (v) compiling and synthesizing findings. Additionally, we will adhere to the Preferred Reporting Items for Systematic Reviews and Meta-Analyses Extension for Scoping Reviews (PRISMA-ScR) (24).

### 2.1 Identifying Research Questions

Following Arksey and O’Malley’s framework (1), we implemented an iterative process to define our research questions. We identified gaps in previous scoping reviews (14,19,20), conducted literature searchers, and engaged in discussions with industry and healthcare professionals focused on promoting metabolic health among older adults. We pose the following questions:

RQ1. How prevalent are DHTs in targeting metabolic disorders in older adults?

RQ2. What specific classes of DHTs have been implemented to address metabolic disorders in older adults?

RQ3. By whom are DHTs used in metabolic disorders contexts?

The formulation of these research questions was guided by the PIO (Population, Intervention, Outcome) concept, as detailed in Table 2, drawing on methodologies from previous studies (22,25).

**Table 2:** The PIO framework for the eligibility of studies.

| Concept | Determinants |
| --- | --- |
| P – Population | Older people (65 or above) |
| I – Intervention | Digital Health Technologies |
| O – Outcome | Metabolic Disorders |

### 2.2 Identifying relevant studies

The methodology for searching literature in this scoping review is outlined in Table 2, based on each PIO (Population, Intervention, Outcome) concept. Initial search terms were established through a preliminary review, particularly by identifying search terms adopted in prior studies on DHTs, older adults and metabolic health (5,21) (See Table 3). To ensure the search was both precise and comprehensive, all authors participated in a consensus-building process. The literature search will span across five major electronic databases: Cochrane, Embase, PubMed, Scopus, and Web of Science, focusing on publications from January 2014 to January 2024, to capture recent advancements in technology. For each database, we will use pre-identified terms on DHTS, metabolic health, and older adults (See Table 3) to search titles, abstracts, and, when available, Medical Subject Headings (MeSH) terms, or else keywords. Within each PIO concept, the Boolean operator ‘OR’ will be used to combine search terms, and then the different concepts will be connected using the ‘AND’ operator.

**Table 3:** Search terms derived for the PIO framework.

| Concept | Search Terms |
| --- | --- |
| 1: Older people (65 or above) (21) | (Older adult* OR Older person OR Elderly OR 65 or above) |
| 2: DHTs (21) | (Internet OR Telemonitor* OR Teleconsultat* OR<br>Telehealth OR Telecare OR Website* OR<br>Apps* OR Application* OR Digital OR<br>eHealth* OR<br>mHealth* OR Health and Wellness* OR Pa-<br>tient Monitoring* OR Care Support* OR Dig-<br>ital Diagnostics* OR Digital Therapeutics*) |
| 3: Metabolic Disorders (5) | ("metabolic health") OR metabolic syn-<br>drome[MeSH Major Topic]) OR hyperglyce-<br>mia[MeSH Major Topic]) OR insulin re-<br>sistance[MeSH Major Topic]) OR diabetes<br>mellitus, type 2[MeSH Major Topic]) OR<br>obesity[MeSH Major Topic]) OR<br>dyslipidemias[MeSH Major Topic]) OR hy-<br>perlipidemias[MeSH Major Topic]) OR hy-<br>pertriglyceridemia[MeSH Major Topic]) OR<br>hypercholesterolemia[MeSH Major Topic]) |
|  | OR hypertension[MeSH Major Topic]) OR non-alcoholic fatty liver disease[MeSH Major Topic] |

*Note:* Search terms adapted from Michels et al. & Cheng et al. (5,21)

### 2.3 Selection of eligible studies

The process of reviewing titles and abstracts will adhere to the PIO framework (outlined in Table 2), using the inclusion and exclusion selection criteria detailed in Table 4 to guarantee that the studies included directly follow our a-priori defined research questions.

**Table 4:** Inclusion and Exclusion Criteria.

| Inclusion Criteria | Exclusion Criteria |
| --- | --- |
| Articles published between January 2014 and January 2024<br>Original or review research articles Qualitative (qualitative descriptive, phenomenological, ethnographical, grounded theory, realistic evaluation, action research, and so on), quantitative (random controlled trials, cohort study, case-control and cross-sectional, and so on) or reviews (systematic review, meta-analysis, scoping review)<br>Articles with a primary focus on DHTs for metabolic health<br>Articles with a primary focus on DHTs for older adults<br>Older individuals 65 years of age or older (or where the average age of the study sample was 65 years of age or older) who had any metabolic disorders (such as insulin resistance, hyperglycemia, type 2 diabetes, obesity, hyperlipidemia, hypertension, non-alcoholic fatty liver disease (NAFLD), and metabolic syndrome) | Articles not in English<br>Articles that do not primarily focus on metabolic syndrome in older adults (aged 65 and above).<br>Articles which discuss metabolic disorders negligibly or with other comorbidities/health conditions<br>Articles that mention DHTs briefly or as an insignificant part<br>Book chapters, commentaries, conference proceedings, editorials, interviews, opinion pieces, proposals, reports, short news, study protocols)<br>Non-human studies |

### 2.4 Charting the data

A form for extracting data will be employed to collect relevant information from every selected article (See Table 5). The process of charting this data will be conducted by hand.

**Table 5:** Data Charting Form adapted from Schneider et al. (2023) (22)

|  |  |
| --- | --- |
| Title of study |  |
| DOI |  |
| Year of publication |  |
| Author name/s |  |
| Author location/s |  |
| Study approach | i.e., qualitative or quantitative or mixed method |
| Type of article | i.e., Case Study, Observational Study, RCT, Review, Trial, Other... |
| Study location | i.e., where the study was carried out |
| DHT Category as defined by DTx Alliance | i.e., Digital Therapeutics, Digital Diagnostics, Care Support, Patient Monitoring, Health and Wellness |
| DHT Use whom | i.e., by who is DHT used (e.g., healthcare provider/researcher/patient) |
| Outcome measured | i.e., the primary outcome being measured in the study |
| Notes |  |

### 2.5 Collating, summarizing, and reporting results

After completing full-text reviews guided by the data extraction charts, we will synthesize the extracted data in descriptive frequency tables. In line with our three research questions, we will first describe the overall frequency of DHTs in studies targeting metabolic disorders in older adults. We will first conduct a comprehensive review of relevant literature and report the prevalence of DHTs across the studies included in our sample (RQ1). Second, we will describe the frequency of each DHT category (promoting health & wellness, (ii) patient monitoring, (iii) care support, (iv) digital diagnostics, and (v) digital therapeutics (15) in targeting metabolic disorders among older adults (RQ2). Next, we will describe by whom are DHTs used across all the six DHTs categories. By categorizing the different types of DHTs, we aim to provide a comprehensive understanding of their utilization in addressing metabolic disorders among older adults and summarize the current DHTs landscape in targeting metabolic disorders.

### 2.6 Ethics and dissemination

The current review will compile information from existing literature and thus does not require additional ethical clearance. Findings from the scoping review will offer potential relevant insights to researchers, policymakers, and healthcare practitioners about the degree to which DHTs are used to target metabolic health orders in older adults and through what relevant approaches, i.e., via health & wellness, patient monitoring, care support, digital diagnostics, and digital therapeutics. The findings will be shared via a scholarly article in a peer-reviewed journal and presented at academic conferences.

## 3 DISCUSSION & CONCLUSION

Overall, the proposed scoping review aims to synthesize the prevalence of DHTs and provide insight into DHTs are used to diagnose, manage, and possibly prevent metabolic disorders in older adults. This scoping review will contribute to a growing body of literature on DHTs in healthcare by specifically focusing on a key demographic of older adults in metabolic disorder contexts. Compared with prior review articles, this study stands out as the first, to our knowledge, to systematically categorize DHTs specifically for older adults, with a concentrated focus on metabolic disorders. To this end, the scoping review presents an important step in standardizing the literature by adopting the DTx Alliance classification framework (8,9). By adopting the DTx Alliance classification framework, we aim to help standardize terminology and categorization within the literature, thereby potentially improving consistency and comparability across studies.

The scoping review presents several significant limitations. As this study encompasses articles featuring a variety of study designs and does not evaluate their quality, it is not designed to resolve questions regarding specific recommendations of DHTs for managing metabolic disorders in older adults. Additionally, the scope of this review is restricted; articles not written in English or lacking a full-text version will be excluded. We also exclude non-peer-reviewed industry sources, which may overlook cutting-edge developments that have not yet been incorporated into the academic literature. Upon completing the full-text review, we intend to comprehensively present both the strengths and limitations of our findings. Furthermore, any deviations from the initial protocol will be transparently reported in the final scoping review.

Overall, our efforts to synthesize the prevalence and use of DHTs in managing metabolic disorders among older adults ae expected to highlight essential areas where DHTs are present versus lacking in the existing literature. Specifically, our findings could reveal key areas where DHTs are underutilized among older adults in metabolic health contexts, thus presenting opportunities for increased DHT inclusion and enhancement. Our results may also offer insights into strategic areas for expanding the use of DHTs for older adults. Together, this review will help better understand the DHTs landscape to tackle metabolic health in the older population.

## Data Availability

All data produced in the present study are available upon reasonable request to the authors

## Footnotes

### Author Statement

All authors have made substantial intellectual contributions to the development of this protocol and its revisions. The search question originated with TK, MJ and PH, then was expanded upon by MB and EF. The methodology and design of the review were conceived by PH, guided by suggestions from TK. The development and refinement of the search terms were a collaborative effort between MJ and PH, incorporating feedback and revisions from TK, MB and EF. The final version of the manuscript received unanimous approval from all authors.

### Patient consent

Not required.

### Provenance and peer review

Not commissioned; externally peer reviewed.

